# Associations between psychiatric disorders, COVID-19 testing probability and COVID-19 testing results: Findings from a population-based study

**DOI:** 10.1101/2020.04.30.20083881

**Authors:** Dennis van der Meer, Justo Pinzón-Espinosa, Bochao D. Lin, Joeri K. Tijdink, Christiaan H. Vinkers, Sinan Guloksuz, Jurjen J. Luykx

**Author notes:** These authors contributed equally (Dennis van der Meer, Justo Pinzón-Espinosa). **Correspondence to:** D van der Meer and J Pinzón-Espinosa.

## Abstract

**Objective:** To compare prevalence of COVID-19 testing and test outcomes among individuals with psychiatric disorders to those without such diagnoses, and to examine the associations of testing probability and outcome with psychiatric diagnosis categories.

**Design:** Large population-based study to perform association analyses of psychiatric disorder diagnoses with COVID-19 testing probability and such test results, by using two-sided Fisher exact tests and logistic regressions.

**Setting:** UK Biobank.

**Participants:** 1 474 men and women of British ancestry that had been tested for COVID-19, with a mean age of 58.2 years.

**Main outcome measures:** COVID-19 testing probability and COVID-19 test results.

**Results:** Individuals with psychiatric disorders were overrepresented among the 1 474 UKB participants with test data: 23% of the COVID-19 test sample had a psychiatric diagnosis compared to 10% in the full cohort (p<0.0001). This overrepresentation persisted for each of the specific psychiatric disorders tested. Furthermore, individuals with a psychiatric disorder (p=0.01), particularly with substance use disorder (p<0.005), had negative test results significantly more often than individuals without psychiatric disorders. Sensitivity analyses confirmed our results.

**Conclusions:** In contrast with our hypotheses, UKB participants with psychiatric disorders have been tested for COVID-19 more frequently than individuals without a psychiatric history, pleading against the notion that limited health care access is preventing them from undergoing testing. Among those tested, test outcomes were more frequently negative for UKB participants with psychiatric disorders than in others, countering arguments that people with psychiatric disorders are particularly prone to contract the virus.

**SUMMARY BOX:** *What is already known on this topic (2–3 sentences):* We searched PubMed using the terms "COVID-19" combined with "mental health", "psychiatric disorder" or "mental illness" for all articles published in any language before April 21^st^, 2020. Two hundred articles were retrieved, most of which related to the Chinese experience when dealing with the pandemic, including the mental health impact of the COVID-19 pandemic on general population mental health and healthcare workers; and on advancing mental healthcare resources in times of crisis. No evidence was found on testing patterns for severe acute respiratory syndromes (e.g. COVID-19, SARS, MERS) or Ebola virus on people with psychiatric disorders.

*What this study adds (2–3 sentences):* We highlight a positive association between psychiatric disorders and the likelihood of being tested for COVID-19, as well as an association between psychiatric disorders and negative results. The results thus counter arguments that patients with psychiatric disorders are suffering from limited health care access preventing them from undergoing testing. Additionally, these are important findings as they carry the potential to reduce stigma: while people in the general population may be concerned that patients with psychiatric disorders do not comply with containment measures and are susceptible to contract COVID-19, our findings may help counter such concerns.

## Introduction

The 2019 Coronavirus disease (COVID-19) caused by the novel SARS-CoV-2 virus strain emerged in Wuhan, China in late 2019 and has since been declared a pandemic.^1^ As of April 21^st^2020, there have been over 2.4 million cases and 163 thousand deaths due to COVID-19 worldwide, where about half correspond to the European region. The UK, the fifth most affected country, has reported over 124 000 cases and over 16 500 deaths.^2^ The challenges of this pandemic to health systems, like the National Health System (NHS) in the UK, include workforce scarcity, insufficient infrastructure and limited testing capacity; these could lead to undetected worsening of long-standing conditions, such as psychiatric disorders.^3^ During epidemics, people with psychiatric disorders may be more susceptible to infections, suffer from complications, and have more difficulties to access health services.^4^ Moreover, during epidemics, people with psychiatric disorders may be at risk for relapse or worsening of their conditions due to higher stress susceptibility compared to the general population as well as due to mobility restrictions possibly preventing them from attending outpatient services.^4^ As a result, innovative approaches to mental health have been increasingly put in place such as telepsychiatry^5,6^ and home hospitalization care.^7^

The aforementioned worries have mobilized mental health professionals to voice concerns about the impact of COVID-19 on people with psychiatric disorders.^8,9^ Due to marginalization, including poor access to health care, and stigma, many psychiatrists are worried that patients with psychiatric and substance use disordersare precluded from receiving timely and appropriate testing.^10^ Second, individuals with a psychiatric disorder may be at increased risk for COVID-19 complications due to comorbid conditions (cardiovascular, respiratory, and metabolic conditions, such as obesity)^11–14^ and potential higher lack of complying with government measures. However, these issues have remained debatable given the current lack of epidemiological data on associations of psychiatric disorders with COVID-19 testing rates and testing outcomes. More research has therefore been called for to address these questions in the current state of pandemic.^15^ To address the questions of whether psychiatric disorders have any association with frequencies of testing and the results of such tests,, we have targeted a large population-based study (the UK Biobank, UKB)^16,17^ to perform association analyses of psychiatric disorder diagnoses with COVID-19 testing probability and such test results. We hypothesized that people with psychiatric disorders are tested for COVID-19 less frequently than people without a psychiatric disorder and that people with psychiatric disorders more frequently test positive than people without psychiatric disorders.

## Methods

The full UKB cohort consists of 502 505 individuals recruited between 2006 and 2010, out of which 157 366 participants have information on a mental health questionnaire.^18^ The composition, set-up, and data gathering protocols of the UKB have been extensively described elsewhere.^19,20^ We made use of data from UKB participants whose COVID-19 test results were released on April 21^st^, 2020 under application access code 55392.^21^ COVID-19 test results in the UKB are mostly derived from samples from nose/throat swabs (or a lower respiratory tract sample in intensive care settings), on which polymerase chain reaction (PCR) is performed. The first data wave released comprises results from March 16^th^, 2020 onwards, as after this date testing in the UK was largely restricted to individuals with symptoms in hospital. Therefore, COVID-19 test results after this date are considered as a surrogate for severe disease.^22^ Before analyses, duplicate entries of test results were removed from the COVID-19 testing results by selecting the latest test results for each participant.

UK Biobank has received ethics approval from the National Health Service National Research Ethics Service (ref 11/NW/0382). We used the STROBE cross sectional reporting guidelines to assess research quality.^23^

For our main analyses we selected International Classification of Diseases, v10 (ICD10) diagnoses from UKB data field 41270.^24^ We compared testing prevalence and testing outcome in individuals with and without a diagnosis of a psychiatric disorder (F codes). To check whether testing prevalence and test results resemble other health conditions, we then included several additional ICD-10 diagnoses in the analyses: 1) individuals with respiratory or cardiovascular diseases as these are particularly at risk for hospitalization following infection with COVID-19, (ICD-10 codes Jxx and 10x-17x); 2) people with metabolic diseases as these are highly prevalent among people with psychiatric disorders and may contribute to much of their generally poor health (codes E0x-E1x and E4x-E7x); and 3) those with central nervous system (CNS) neurological disorders, as a comparison category of diseases resembling psychiatric disorders with regards to symptomatology and hypothesized neurobiological underpinnings (G0x-G4x).

Subsequently, we investigated each of the major psychiatric disorder categories with a prevalence above a threshold of 5% (n=74) among individuals within the subsample that had test results available. We therefore included substance use disorders (F1x), mood disorders (F3x), and anxiety disorders (F40-F41) in the analyses.

All data was analysed in R v3.6.1.^25^ We applied two statistical tests to answer the following primary research questions:

1. Are people with a psychiatric disorder more or less likely to undergo COVID-19 testing than people without such a diagnosis? To answer this first question, we used two-sided Fisher’s exact tests to examine distributions of individuals tested compared to the full UKB cohort.
2. Are people with a psychiatric disorder more or less likely to test positive for COVID-19 compared to those without such a diagnosis? To answer this second question, we ran logistic regression models, using the COVID-19 test results as a dichotomous outcome (negative/positive), with the ICD-10 diagnoses or mental health categories as predictors. We report the change in log odds (ß) from these models.

Age, sex, body mass index (BMI), and assessment centre were used as covariates in the logistic regression models as the first three have been associated with both psychiatric disorders’ and COVID-19 prevalences, while assessment centre was added to these models to prevent regional differences impacting the results. To assess the robustness of our findings, we further ran a sensitivity analysis additionally covarying for social-economic status (SES), as measured through the Townsend deprivation index, which has previously been used in the UKB,^26,27^ and pre-existing cardiovascular, respiratory, and metabolic conditions as those are commonly observed in people with psychiatric disorders.

Secondary analyses included population-level information on mental health based on mental health questionnaire items asking participants whether they had ever experienced a core symptom of the major mental health categories (data category 136). For example, the two questions on depression were "Have you ever had prolonged feelings of sadness or depression?" and "Have you ever had prolonged loss of interest in normal activities?", tapping into the two core symptoms of major depressive disorder of depressed mood and anhedonia.^28^ If participants had an affirmative response to either of these items, we scored the depression mental health category as present, otherwise as absent. This way, we aimed to examine relationships of the presence vs absence of mental health symptoms (depressive, manic, anxiety, addiction, psychotic experiences, self-harm, and happiness items) with both testing probability and testing results in this general population cohort. To test this, we used identical analysis approaches as for the primary analyses, i.e. Fisher’s exact tests and logistic regression with the same covariates as mentioned above. We also ran a sensitivity analysis for this secondary analysis by including the abovementioned additional covariates, similar to the primary analyses. We ran these analyses with and without including individuals with a diagnosis of psychiatric disorder to disentangle whether people with any such diagnosis are driving results, and to what extent continuous measures of mental health are associated with COVID-19 testing probability and outcome. Please see the supplementary information for an overview of all mental health items and more information on these analyses.

## Results

The UKB subsample with COVID-19 test results available consisted of 1 474 unique individuals. Of these, 842 tested negative (57.1%) and 632 tested positive (42.9%) for the virus. Individuals tested were significantly older than untested UKB participants (58.2 years (standard deviation (SD) 8.8) vs 57.0 years (SD) 8.1; p=2.2*10^−7^), there were more men among those tested (54.4%) than among those not tested (46.6%), p=2.1*10^−9^, and the average Townsend deprivation index was lower among those tested than among those not tested (−0.16 (SD 3.53) vs −1.30 (SD 3.09), p<1*10^−16^). There were no significant differences in age (p=.51), sex (p=.14) or SES (p=.13) between those testing positive versus negative.

We found that individuals with a psychiatric disorder were overrepresented among those tested, making up 23% of this sample compared to 10% in the full UKB cohort (p<0.0001; Table 1). This overrepresentation was similar to, or even higher than, that of people with diagnoses of cardiovascular, respiratory, metabolic, or neurological conditions (Table 1). Furthermore, this overrepresentation was also present for each of the specific psychiatric disorder categories investigated (Table 1).

**Table 1.**
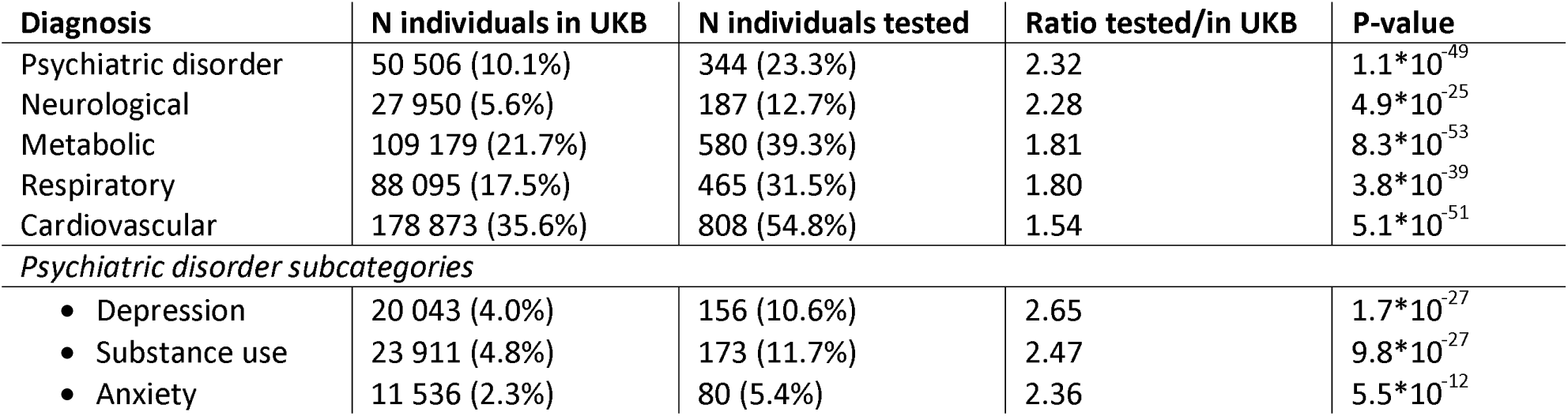
*Comparison of number of individuals in the full UKB cohort with those among the COVID-19 tested subset, per diagnostic group, ordered by decreasing ratio. The columns indicate the number of individuals with a specific diagnosis in either the full UKB cohort or in the tested subset, and the resulting ratio. The numbers in brackets indicate the corresponding percentage of individuals. The p-value is determined by Fisher’s exact test*.

Among those tested, individuals with a diagnosis of a psychiatric disorder significantly less frequently tested positive COVID-19 test compared to those without such a diagnosis (p=0.01, ß=-0.35; Figure 1A). When looking into specific psychiatric disorders, we found that particularly individuals with substance use disorders were significantly less likely to test positive (p=0.0002;, ß=-0.70; Figure 1B).While people with anxiety and depressive disorders were also less likely to test positive than those without such a diagnosis, those results were non-significant. The pattern of results did not change in our sensitivity analysis where we additionally co-varied for pre-existing cardiovascular, respiratory, and metabolic conditions, as well as SES; both general psychiatric disorder and substance use specifically remained significantly associated with test outcome (Figure S1).

**Figure 1.**
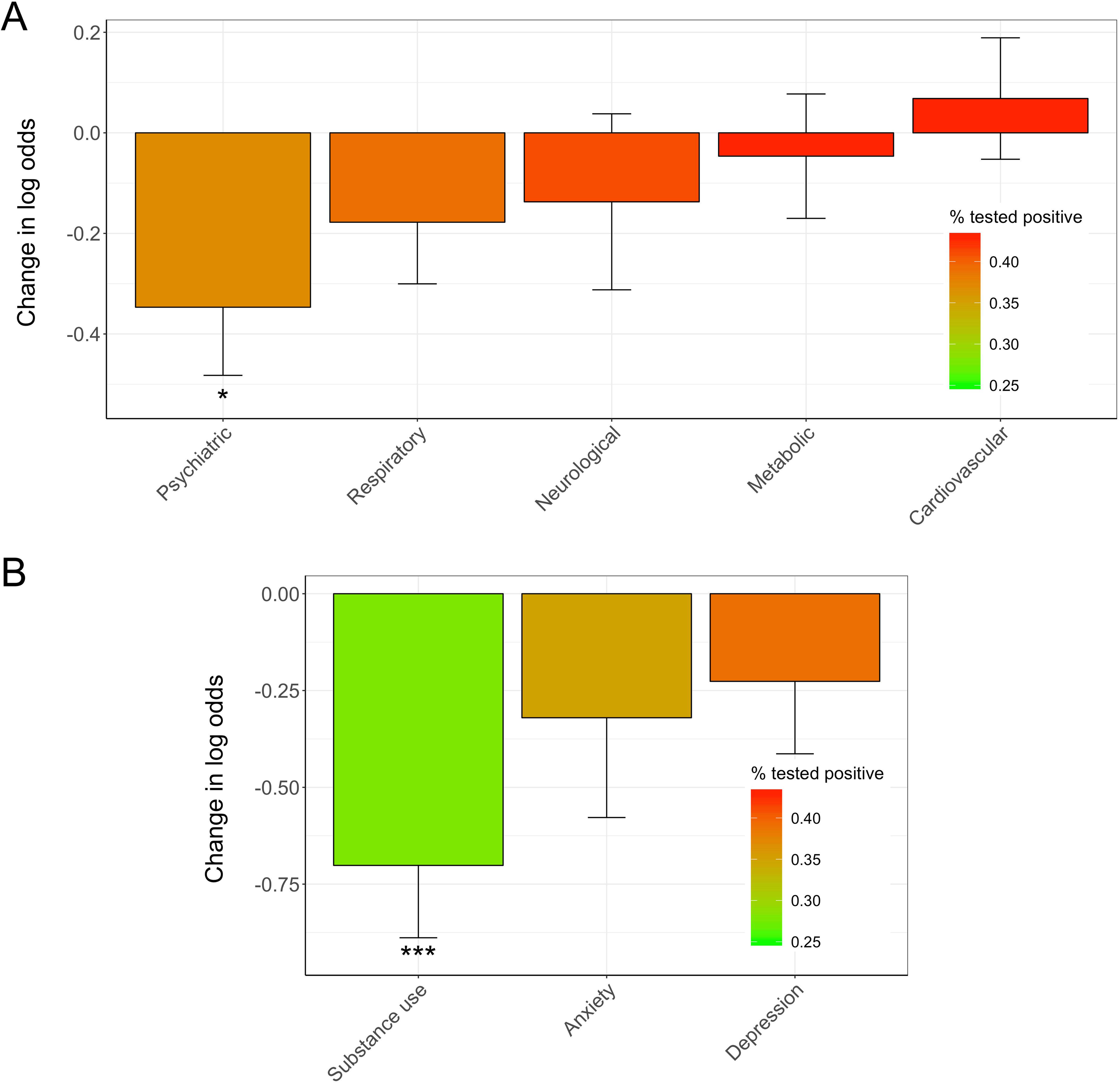
Bar plot of change in log odds for testing positive, by ICD10 diagnosis. Change in log odds is shown on the y-axis, on the x-axis are the diagnoses, colors indicate percent that tested positive. Superscript indicate significance: * p<·05, ** p<·005, *** p<·0005. A) shows the results for the main ICD10 diagnoses, B) those for the psychiatric disorders subcategories.

After removing individuals with a psychiatric disorder from the sample, there were no statistically significant associations between responses to the mental health items and test frequency or test results (see supplemental information for the full results).

## Discussion

Contrary to our hypotheses, we found that individuals with psychiatric disorders have been more frequently tested for COVID-19 compared to those without a diagnosis. Furthermore, among those tested, individuals with psychiatric disorders, substance use disorders in particular, had lower odds of testing positive than individuals without such a diagnosis. We believe these are important findings as they carry the potential to reduce stigma: while people in the general population may be concerned that individuals with psychiatric disorders do not comply with containment measures and are thus susceptible to contract COVID-19, our findings may help counter such concerns. Our findings also may help diminish concerns over limited health care access preventing people with psychiatric disorders from undergoing testing.

Possibly, people with psychiatric disorders are being tested more frequently because of comorbid conditions, higher levels of anxiety about contracting COVID-19 or perhaps a combination of both. Referring physicians’ concerns with COVID-19 in people with psychiatric disorders may contribute to relatively high testing rates. Such reasoning may also, at least in part, explain higher rates of negative COVID-19 test results in people with psychiatric disorders. Additional underlying reasons for the latter may be that they live relatively more socially isolated than people without such diagnoses. A non-controlled study has previously shown people with psychosis and mood disorders have about 1.7 social contacts in a week outside home, workplace or healthcare setting, and moderate feelings of loneliness.^29^ Furthermore, a study on the relationship between living alone and psychiatric disorders using the 1993, 2000, and 2007 National Psychiatric Morbidity Surveys in the United Kingdom found a positive association between both variables that was up to 84% explained by loneliness.^30^ Among young adults in modern Britain, relatively lonely individuals have been shown to be more likely to suffer from depressive, anxiety, and alcohol use disorders.^31^ Thus, lonelier, more socially isolated people such as those with psychiatric disorders may normally, and now even more so, be in confinement and have less social contact than people without psychiatric disorders, reducing the likelihood of testing positive for COVID-19.

Furthermore, when looking at the symptom-level in the UK Biobank sample (mental health questionnaire items), no relation was found between test prevalence or outcome and continuous measures of depressive, manic, anxiety, addiction, psychotic experiences, self-harm or happiness items in those without past or current diagnosis of psychiatric disorder. Therefore, clinical cases of psychiatric disorders, and not sub-syndromic individuals, appear to drive our primary findings. Statistical power may currently hamper these analyses.

Limitations of this study include the relatively small sample size, the fact that the UKB is not fully representative of the general population,^32–34^ and absence of replication in other cohorts. The small sample size precluded individuals with diagnosis of severe psychiatric disorders, like schizophrenia, schizoaffective disorder, or bipolar disorders, to be represented in the analyses. Furthermore, assessment centre was used as a proxy for geographical location, and this variable was set at the start of recruitment, e.g. if individuals moved after the initial assessment, this was not possible to take into consideration. Yet, two preliminary conclusions can be drawn based on the current dataset given the convergence of findings for a range of psychiatric disorders and similarities between testing probabilities. First, individuals with a psychiatric disorder are not less likely to undergo testing for COVID-19 than those without psychiatric disorders. Second, patients with psychiatric disorders do not test positive more frequently than people undergoing testing without such conditions. We encourage other researchers to perform similar analyses in other cohorts, as well as further research when more data from the UK Biobank become available, e.g. into associations between extended psychiatric symptom-level data, COVID-19 symptom severity and mortality.

## Data Availability

This research has been conducted using the UK Biobank Resource under Application Number 55392.

## Contributors

All authors conceived the study and interpreted the results. DvdM analysed the data. JPE, DvdM, and JL wrote the manuscript, which was revised by BL, JT, CH, and SG. JPE and DvdM are the guarantors.

## Transparency Statement

The lead authors (the manuscript’s guarantors) affirm that this manuscript is an honest, accurate, and transparent account of the study being reported; that no important aspects of the study have been omitted; and that any discrepancies from the study as planned (and, if relevant, registered) have been explained.

## Data Sharing

The data reported in this paper are available via application directly to the UK Biobank.

## Patient Involvement

This study was conducted using the UK Biobank resource. Details of patient and public involvement in the UK Biobank are available online (http://www.ukbiobank.ac.uk/about-biobank-uk/). No patients were specifically involved in setting the research question or the outcome measures, nor were they involved in developing plans for recruitment, design or implementation of this study. No patients were asked to advise on data interpretation or writing up of results. There are no specific plans to disseminate the results of the research to study participants, but the UK Biobank disemminates key findings from projects on its website.

## Copyright Statement

The Corresponding Authors have the right to grant on behalf of all authors and do grant on behalf of all authors, <u>a worldwide licence</u> to the Publishers and its licensees in perpetuity, in all forms, formats and media (whether known now or created in the future), to i) publish, reproduce, distribute, display and store the Contribution, ii) translate the Contribution into other languages, create adaptations, reprints, include within collections and create summaries, extracts and/or, abstracts of the Contribution, iii) create any other derivative work(s) based on the Contribution, iv) to exploit all subsidiary rights in the Contribution, v) the inclusion of electronic links from the Contribution to third party material where-ever it may be located; and, vi) licence any third party to do any or all of the above.

## Acknowledgments

The authors are extremely grateful to the participants who took, and continue taking, part in the UK Biobank cohort study. This research has been conducted using the UK Biobank Resource under Application Number 55392. The authors also thank Tobias Kaufmann for his input to our analyses and manuscript.

## REFERENCES

1 Wang C, Horby PW, Hayden FG, Gao GF. A novel coronavirus outbreak of global health concern. Lancet 2020; 395: 470–3.

2 World Health Organization. WHO COVID-19 Dashboard. 2020. https://who.sprinklr.com/ (accessed April 22, 2020).

3 Willan J, King AJ, Jeffery K, Bienz N. Challenges for NHS hospitals during covid-19 epidemic. BMJ. 2020; 368. DOI:10.1136/bmj.mlll7.

4 Yao H, Chen JH, Xu YF. Patients with mental health disorders in the COVID-19 epidemic. The Lancet Psychiatry 2020; 7: e21.

5 Zhou X, Snoswell CL, Harding LE, et al. The Role of Telehealth in Reducing the Mental Health Burden from COVID-19. Telemed e-Health 2020; 26: tmj.2020.0068.

6 Liu S, Yang L, Zhang C, et al. Online mental health services in China during the COVID-19 outbreak. The Lancet Psychiatry 2020; 7: e17–8.

7 Garriga M, Agasi I, Fedida E, et al. The role of Mental Health Home Hospitalization Care during the COVID-19 pandemic. Acta Psychiatr Scand 2020; 2405: 0–2.

8 Torales J, O’Higgins M, Castaldelli-Maia JM, Ventriglio A. The outbreak of COVID-19 coronavirus and its impact on global mental health. Int J Soc Psychiatry 2020;: 002076402091521.

9 Luykx JJ, Vinkers CH, Tijdink JK. Psychiatry in times of COVID-19: an imperative for psychiatrists to act now. JAMA Psychiatry 2020; **In press**.

10 Druss BG. Addressing the COVID-19 Pandemic in Populations With Serious Mental Illness. JAMA Psychiatry 2020; 2019: 2019–20.

11 Simonnet A, Chetboun M, Poissy J, et al. High prevalence of obesity in severe acute respiratory syndrome coronavirus-2 (SARS-CoV-2) requiring invasive mechanical ventilation. Obesity (Silver Spring) 2020; published online April 9. DOI:10.1002/oby.22831.

12 Sattar N, Mclnnes IB, McMurray JJ V. Obesity a Risk Factor for Severe COVID-19 Infection: Multiple Potential Mechanisms. Circulation 2020;: CIRCULATIONAHA.120.047659.

13 Dietz W, Santos-Burgoa C. Obesity and its Implications for COVID-19 Mortality. Obesity 2020; published online April 1. DOl:10.1002/oby.22818.

14 Rubino S, Kelvin N, Bermejo-Martin JF, Kelvin D. As COVID-19 cases, deaths and fatality rates surge in Italy, underlying causes require investigation. J Infect Dev Ctries 2020; 14: 265–7.

15 Holmes EA, O’Connor RC, Perry VH, et al. Multidisciplinary research priorities for the COVID-19 pandemic: a call for action for mental health science. The lancet Psychiatry 2020; published online April. DOI:10.1016/S2215-0366(20)30168-1.

16 Bycroft C, Freeman C, Petkova D, et al. The UK Biobank resource with deep phenotyping and genomic data. Nature 2018; 562: 203–9.

17 UK BIOBANK MAKES INFECTION AND HEALTH DATA AVAILABLE TO TACKLE COVID-19 | UK Biobank. 2020. https://www.ukbiobank.ac.uk/2020/04/covid/ (accessed April 22, 2020).

18 Davis KAS, Coleman JRI, Adams M, et al. Mental health in UK Biobank—development, implementation and results from an online questionnaire completed by 157 366 participants: a reanalysis. BJPsych Open 2020; 6. DOI:10.1192/bjo.2019.100.

19 Sudlow C, Gallacher J, Allen N, et al. UK Biobank: An Open Access Resource for Identifying the Causes of a Wide Range of Complex Diseases of Middle and Old Age. PLoS Med 2015; 12: e1001779.

20 Miller KL, Alfaro-Almagro F, Bangerter NK, et al. Multimodal population brain imaging in the UK Biobank prospective epidemiological study. Nat Neurosci 2016; 19:1523–36.

21 Enhancing resilience in psychosis through within and between-family polygenic risk scoring, Genex Gene interactions and gene-environment (GxE) prediction models (REGENESIS) | UK Biobank. https://www.ukbiobank.ac.uk/2019/ll/enhancing-resilience-in-psychosis-through-within-and-between-family-polygenic-risk-scoring-gene-x-gene-interactions-and-gene-environment-gxe-prediction-models-regenesis/ (accessed April 22, 2020).

22 UKB: Data-Field 40100. 2020. http://biobank.ndph.ox.ac.uk/showcase/field.cgi?id=40100 (accessed April 30, 2020).

23 von Elm E, Altman DG, Egger M, Pocock SJ, Gøtzsche PC, Vandenbroucke JP. The Strengthening the Reporting of Observational Studies in Epidemiology (STROBE) statevon Elm, Erik, Douglas G. Altman, Matthias Egger, Stuart J. Pocock, Peter C. Gøtzsche, and Jan P. Vandenbroucke. 2008. “The Strengthening the Reporting of Observational. J Clin Epidemiol 2008; 61: 344–9.

24 World Health Organization. International Classification of Diseases (ICD-10). 2010. https://icd.who.int/browsel0/2010/en (accessed April 21, 2020).

25 R Core Team. R: A language and environment for statistical computing. 2019. https://www.r-project.org/.

26 Tyrrell J, Jones SE, Beaumont R, et al. Height, body mass index, and socioeconomic status: Mendelian randomisation study in UK Biobank. BMJ 2016; 352. DOI:10.1136/bmj.i582.

27 Townsend P, Phillimore P, Beattie A. Health and deprivation: inequality and the North. Routledge, 1988.

28 Malhi GS, Mann JJ. Depression. Lancet. 2018; 392: 2299–312.

29 Giacco D, Palumbo C, Strappelli N, Catapano F, Priebe S. Social contacts and loneliness in people with psychotic and mood disorders. Compr Psychiatry 2016; 66: 59–66.

30 Jacob L, Haro JM, Koyanagi A. Relationship between living alone and common mental disorders in the 1993, 2000 and 2007 National Psychiatric Morbidity Surveys. PLoS One 2019; 14. DOI:10.1371/journal.pone.0215182.

31 Matthews T, Danese A, Caspi A, et al. Lonely young adults in modern Britain: Findings from an epidemiological cohort study. Psychol Med 2019; 49: 268–77.

32 Keyes KM, Westreich D. UK Biobank, big data, and the consequences of non-representativeness. Lancet. 2019; 393: 1297.

33 Fry A, Littlejohns TJ, Sudlow C, et al. Comparison of Sociodemographic and Health-Related Characteristics of UK Biobank Participants with Those of the General Population. Am J Epidemiol 2017;186: 1026–34.

34 Abdellaoui A. Regional differences in reported Covid-19 cases show genetic correlations with higher socio-economic status and better health, potentially confounding studies on the genetics of disease susceptibility. *medRxiv* 2020;: 2020.04.24.20075333.

